# Biobank scale pharmacogenomics informs the genetic underpinnings of simvastatin use

**DOI:** 10.1101/2020.11.23.20235846

**Authors:** Frank R Wendt, Dora Koller, Gita A Pathak, Daniel Jacoby, Edward J Miller, Renato Polimanti

## Abstract

**Background and Purpose:** Studying drug metabolizing enzymes, encoded by pharmacogenes (PGx), may inform biological mechanisms underlying the diseases for which a medication is prescribed. Until recently, PGx loci could not be studied at biobank scale. Here we analyze PGx haplotype variation to detect associations with medication use in the UK Biobank.

**Methods:** In 7,649 unrelated African-ancestry (AFR) and 326,214 unrelated European-ancestry (EUR) participants from the UK Biobank, aged 37-73 at time of recruitment, we associated clinically-relevant PGx haplotypes with 265 (EUR) and 17 (AFR) medication use phenotypes using generalized linear models covaried with sex, age, age^2^, sex×age, sex×age^2^, and ten principal components of ancestry. Haplotypes across 50 genes were assigned with Stargazer. Our analyses focused on the association of PGx haplotype dose (quantitative predictor), diplotype (categorical predictor), and rare haplotype burden on medication use.

**Results:** In EUR, *NAT2* metabolizer phenotype (OR=1.05, 95% CI: 1.03-1.08, p=7.03×10^−6^) and activity score (OR=1.09, 95% CI: 1.05-1.14, p=2.46×10^−6^) were associated with simvastatin use. The dose of N-acetyltransferase 2 (*NAT2*)*1 was associated with simvastatin use relative to *NAT2*\*5 (*NAT2*\*1 OR=1.04, 95% CI=1.03-1.07, p=1.37×10^−5^) and was robust to effects of low-density lipoprotein cholesterol (LDL-C) concentration (*NAT2*\*1 given LDL-C concentration: OR=1.07, 95% CI=1.05-1.09, p=1.14×10^−8^) and polygenic risk for LDL-C concentration (*NAT2*\*1 given LDL-C PRS: OR=1.09, 95% CI=1.04-1.14, p=2.26×10^−4^). Interactive effects between *NAT2*\*1, simvastatin use, and LDL-C concentration (OR: 0.957, 95% CI=0.916-0.998, p=0.045) were replicated in eMERGE PGx cohort (OR: 0.987, 95% CI: 0.976-0.998, p=0.029).

**Conclusions and relevance:** We used biobank-scale data to uncover and replicate a novel association between *NAT2* locus variation (and suggestive evidence with several other genes) and better response to simvastatin (and other statins) therapy. The presence of *NAT2*\*1 versus *NAT2*\*5 may therefore be useful for making clinically informative decisions regarding the potential benefit (e.g., absolute risk reduction) in LDL-C concentration prior to statin treatment.

**Subject terms:** genetics, genetic association studies, cardiovascular disease

## Introduction

More than half of phase II and III clinical trials fail because of the lack of efficacy of the investigational drug(s).^1^ Predicting drug efficacy in a given disorder is of great clinical interest but given the highly polymorphic nature of many pharmacogenes (PGx) and the heterogeneous nature of drug metabolism, identifying clinically informative genetic variation often requires extremely large sample sizes.^2-7^ Motivated by the hypothesis that studying medication use could reveal additional information about biological mechanisms underlying complex disease, a recent genome-wide association study (GWAS) identified 505 independent loci associated with various medication endorsements in 318,177 subjects from the UK Biobank (UKB), suggesting possible biological mechanisms, drug targets, and expected adverse events.^6^ Among the medication-associated genes discovered, the study found 16 genes already known as therapeutic targets.^6^ A major limitation to using GWAS discoveries to understand PGx risk factors stems from deployment feasibility. At the SNP-level, even polygenic risk scores (PRS; the weighted or unweighted additive genetic risk for a phenotype) explain relatively little phenotypic variation making it difficult to translate SNP-based to clinical practice.^8^

The response to several drugs can be predicted by genetic variants that influence their pharmacokinetics (metabolism) and/or pharmacodynamics (mechanism of action).^9^ Currently 66 Very Important Pharmacogenes (VIP) are registered in the Pharmacogenomics Knowledge Base based on recommendations from the U.S. Food and Drug Administration (FDA) and the Clinical Pharmacogenetic Implementation Consortium.^10, 11^ These recommendations are based on an individual’s diplotype at certain loci – the combination of multi-variant haplotypes termed star (*) alleles.^12^ For instance, the FDA and the European Medicines Agency recommend dose adjustments of amifampridine based on *N-acetyltransferase 2* (*NAT2*) poor metabolizer status. Poor metabolizers should be administered no more than 15 mg daily dose of amifampridine, a drug for Lambert-Eaton myasthenic syndrome, in divided doses to avoid toxicity.^13^ Because cholesterol lowering statins are used by up to 28% of the adult population^14^, this class of medication has garnered considerable attention with respect to genetically predicted toxicity.^15^ One of the most convincing and clinically informative pieces of evidence for adverse statin effects is increased risk of simvastatin induced myopathy (muscle weakness) in patients carrying the rs4149056-C allele within *solute carrier organic anion transporter family member 1B1* (*SLCO1B1*).^15^

In this study we employ a hypothesis-generating PGx-wide association approach to discover risk loci associated with medication use in European and African ancestry individuals from the UK Biobank. We demonstrate that common haplotype variation in *NAT2* interacts with simvastatin use to produce clinically useful relative decreases in LDL-C concentrations (∼1 mmol/L), of which *NAT2* contributes approximately 1-5%. These data provide clinically informative and actionable associations with PGx variation and the use of hundreds of medications in two ancestries that can be used to design studies characterizing medication-PGx relationships as prognostic indicators of adverse and beneficial outcomes across the human phenome.

## Materials and Methods

### UK Biobank description and participant inclusion

The UK Biobank (UKB) is a large population cohort comprised of >500,000 participants. UKB participants range in age from 37-73 years at time of recruitment and represent a general population sampling of the United Kingdom with no enrichment for specific disorders. A total of 372,854 UKB participants completed a verbal interview by a trained nurse regarding prescription medication use (UKB Field ID 20003). Responses of non-medication use were confirmed by two rounds of nurse-administered interview. Medication endorsements used here reflect regular (weekly or monthly) use of a specific medication rather than short-term medication use (e.g., 1-week course of use; see http://biobank.ndph.ox.ac.uk/showcase/label.cgi?id=100075). This study focused on sex-combined analyses and therefore, oral contraceptives were removed. Based on results from **Power analyses**, we analyzed a total of 265 (EUR) and 17 (AFR) prescriptions. Their use prevalence is provided in Tables S1 and S2 for EUR and AFR ancestries, respectively.

This study was conducted using publicly available previously collected and deidentified genetic data and is exempt from Yale institutional review board approval. The UK Biobank obtained Research Tissue Bank (RTB) approval from its ethics committee that covers the present uses of the resource.

### Very important pharmacogene haplotype calling

Variant call files (VCF) were generated from UKB plink binary files for genomic regions (build hg19) of interest for each pharmacogene (Table S3). Using VCFs as standard input, haplotypes for each individual in the UKB were assigned using Stargazer_v1.0.8 (February 25, 2020 release).^10, 11^ Briefly, Stargazer calls PGx haplotypes using variants (single nucleotide polymorphism, insertion-deletion, and structural variations) recognized by the Pharmacogene Variation Consortium (PharmVar). Beagle was used to phase heterozygous variants using the 1000 Genomes Project reference population. Phased UKB haplotypes were matched to star (*) allele reference haplotypes using a translation table built from publicly available PharmVar and 1000 Genomes Project data. Diplotypes are defined as the combination of haplotypes at a given gene – the relationship between haplotype and diplotype is analogous to the relationship between SNP and genotype. Haplotypes and diplotypes were assigned activity scores (AS) which translate PGx information into qualitative measures of protein/enzyme activity.^12, 14, 16^ Activity scores are then converted to categorical metabolizer phenotypes (MP; e.g., poor, intermediate, normal, rapid metabolism, among others).^10,11^

### Power analyses

We performed power analysis to determine the appropriate sample size (i.e., required number of cases per medication use phenotype) and PGx haplotype frequency appropriate for testing association between the two in EUR and AFR ancestries. Power was assessed with the genpwr^17^ R package for AFR and EUR ancestry cohorts separately due to large differences in sample size between them. We assumed a relatively large effect size distribution (OR=1.25, 1.5, and 2) across three medication use case proportions (EUR = 0.05%, 0.075%, and 0.1%; AFR = 1%, 1.5%, and 2.5%) and minor haplotype frequencies ranging from 1-20% (applied to EUR and AFR). These tests are shown graphically in Figure S1. In EUR we achieved 80% power to detect moderate effect sizes at a case prevalence of 0.1% (325 cases) for haplotypes at 1% frequency. In AFR we selected phenotypes with 2.5% case prevalence (193 cases) and haplotypes observed in at least 5% of the population.

### Association analysis

Generalized linear models were used to associate each medication use phenotype two ways with sex, age, age^2^, sex×age, sex×age^2^, and ten principal components of ancestry as covariates. First, medication use was associated with AS and MP and for each significant association we then evaluated the effects dose of each common PGx haplotype relative to the most common haplotype. Second, for each significant association between haplotype dose and medication use, we compared homozygous reference, heterozygous, and homozygous alternate diplotypes as categorical predictors of medication use. Analysis was performed in R version 3.5.1 using the “binary” family of the glm() function.

### Calculating polygenic risk scores

Polygenic risk scores (PRS) were calculated for each UKB participant using weights of genome-wide significant SNPs from large GWAS of each traits of interest. For SNP effect weights associated with low density lipoprotein cholesterol (LDL-C) concentration, we used 212 LD-independent SNPs that reached genome-wide significance in a study of 94,595 EUR ancestry individuals naïve to lipid lowering medications.^18^ Sample overlap is known to confound PRS calculation so although this base GWAS was not the largest available for LDL-C,^19^ it is the largest, to our knowledge, that lacks sample overlap with the UK Biobank. PRS per UKB participant were calculated using PRSice v2^20^ and the following parameters: clumb-kb = 250, clump-p = 1, clump-r^2^ = 0.1. Note that PRSice v2 was deployed only to calculate PRS for UKB participants and was not used to regress against a target phenotype.

### Statistical considerations

False discovery rates were applied to all analyses based on the number of gene-phenotype, MP-phenotype, or AS-phenotype combinations tested. Note that for selecting genes based on their MP and AS association with medication use, multiple testing correction was applied to AS and MP results independently. FDR multiple testing correction was applied using the p.adjust() function in R.

### Replication

The eMERGE-PGx (Electronic Medical Records and Genomics Pharmacogenetic Sequencing Pilot)^21^ project was used for replication (dbGaP study accession phs000906.v1.p1). Study sites include Children’s Hospital of Philadelphia, Cincinnati Children’s Hospital Medical Center/Boston’s Children’s Hospital, Geisinger Health/University of Washington, Essentia Institute of Rural Health/Marshfield Clinic/Pennsylvania State University, Mayo Clinic, Northwestern University, and Vanderbilt University. Participants (total N=9,010) were recruited to the eMERGE PGx pilot due to either self-reported or algorithm-predicted use of clopidogrel, warfarin, and/or simvastatin.

Due to the target sequencing (PGx regions only) nature of eMERGE PGx Sequencing Pilot, we did not perform genetic ancestry assignment but instead selected all participants who self-identified as non-Hispanic white. For each individual in eMERGE with multiple instances of laboratory measures we retained only the earliest laboratory measure (i.e., the measure made at the individual’s youngest age). To best represent the age distribution of the UKB discovery cohort, the eMERGE cohort (N=2,826; mean age=53.3, standard deviation=12.2) was restricted to the same age range as the UKB EUR (mean age=56.6, standard deviation=8.02, p_diff_=0.873). LDL-C concentration units were normalized across studies using the conversion factor 1 mg/dL = 0.0259 mmol/L.^22^

## Results

### Cohort description

The UK Biobank is a population-level general sampling of >502,000 participants ranging from 37-73 years at the time of recruitment. For each indication of a given medication, the participant was considered a case while lack of use for a given medication qualified a participant as a control. We stratified the UK Biobank participants into unrelated European-descent British (N=326,214) and African-ancestry (N=7,649) cohorts.

### Phenotype associations

Using Stargazer v1.0.8 (February 2020 Release),^10, 11^ 262 star (*) alleles were called across 50 PGx loci in UKB AFR and EUR cohorts (Figure 1 and Table S4). Twenty-seven (EUR) and 25 (AFR) PGx loci had haplotypic variation with known effects on protein function based on data from the PharmVar (Figure S2 and Table S5). Genetically predicted ASs and MPs for each gene were associated with suitably powered medication use phenotypes (265 in EUR and 17 in AFR; see **Methods**) in the UKB with generalized linear models covaried with sex, age, age^2^, sex×age, sex×age^2^, and ten principal components of ancestry. In EUR and AFR, respectively, we detected 279 and 13 nominally significant associations between medication use and PGx MP and AS (Figures 2 and S3 and Tables S6 and S7). After multiple testing correction (N=7,115 each in MP and AS), EUR *NAT2* MP (MP OR=1.05, 95% CI: 1.03-1.08, p=7.03×10^−6^) and AS were associated with simvastatin use (AS OR=1.09, 95% CI: 1.05-1.14, p=2.46×10^−6^). Simvastatin use was nominally associated with seven other PGx loci (Table 1 and Table S6): *CYP2D6* (increased odds), *CYP2C8* (decreased odds), *POR* (decreased odds), *SLCO1B1* (increased odds), *CYP3A5* (decreased odds), *IFNL3* (decreased odds), and *CYP2C9* (decreased odds). In the following sections we characterize the effects of *NAT2* *-allele dose on simvastatin use. In the Supplementary Results we provide two additional sets of results: (1) four PGx-medication use tests that were significant after analysis-wide multiple testing correction – these results survived multiple testing corrected in MP *or* AS tests but not both and (2) EUR and AFR ancestry results meeting a nominal significance threshold (Tables S8-S10).

**Table 1.**
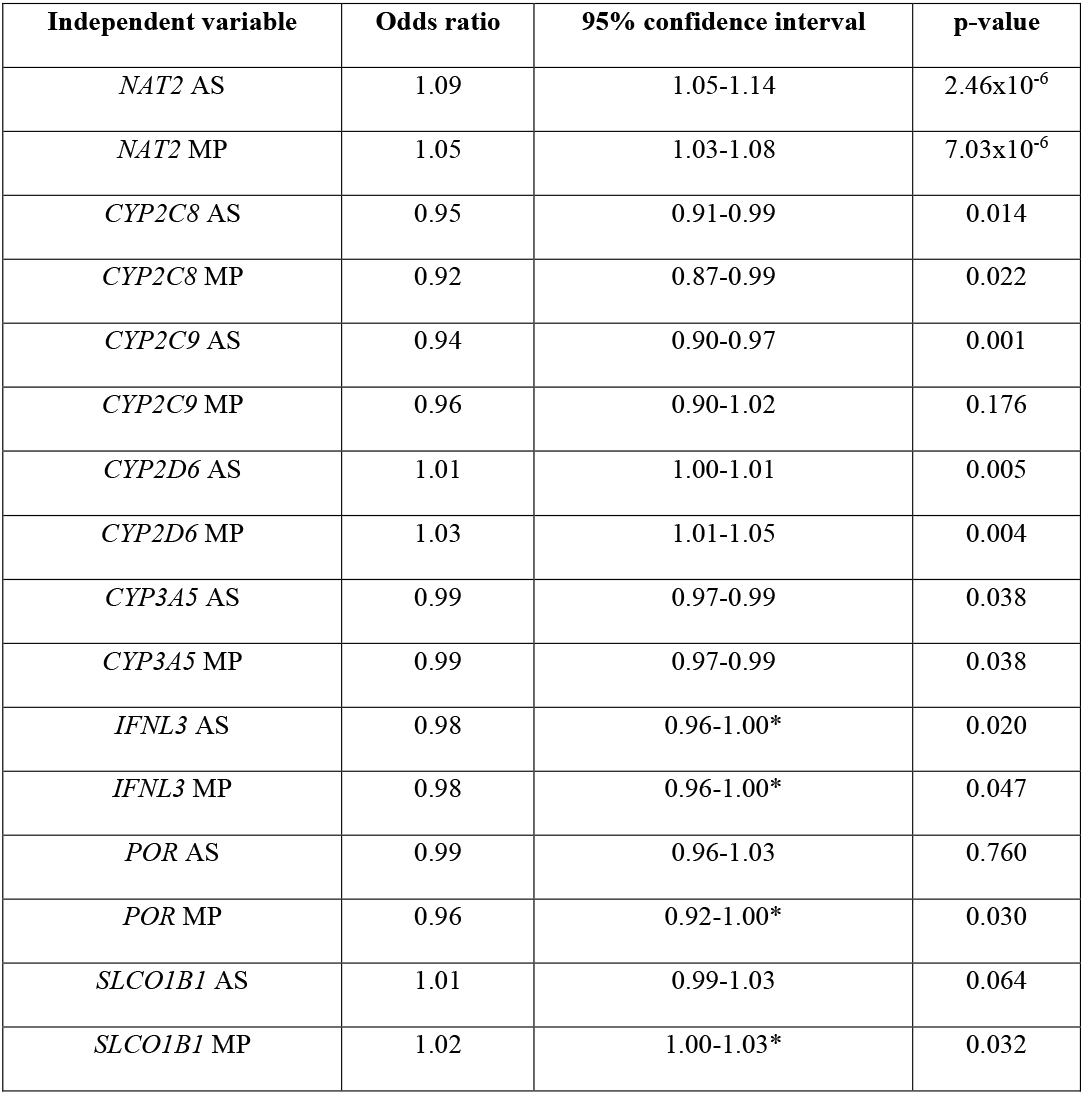
Pharmacogenes (PGx) with evidence of effect on simvastatin use in European ancestry participants from the UK Biobank. Note that at *CYP3A5*, activity score (AS) and metabolizer phenotype (MP) capture the same level of variation (see Figure 2 and Table S6). Asterisks (*) indicate confidence intervals that include OR=1 due to rounding.

| Independent variable | Odds ratio | 95% confidence interval | p-value |
| --- | --- | --- | --- |
| *NAT2* AS | 1.09 | 1.05-1.14 | 2.46x10<sup>-6</sup> |
| *NAT2* MP | 1.05 | 1.03-1.08 | 7.03x10<sup>-6</sup> |
| *CYP2C8* AS | 0.95 | 0.91-0.99 | 0.014 |
| *CYP2C8* MP | 0.92 | 0.87-0.99 | 0.022 |
| *CYP2C9* AS | 0.94 | 0.90-0.97 | 0.001 |
| *CYP2C9* MP | 0.96 | 0.90-1.02 | 0.176 |
| *CYP2D6* AS | 1.01 | 1.00-1.01 | 0.005 |
| *CYP2D6* MP | 1.03 | 1.01-1.05 | 0.004 |
| *CYP3A5* AS | 0.99 | 0.97-0.99 | 0.038 |
| *CYP3A5* MP | 0.99 | 0.97-0.99 | 0.038 |
| *IFNL3* AS | 0.98 | 0.96-1.00* | 0.020 |
| *IFNL3* MP | 0.98 | 0.96-1.00* | 0.047 |
| *POR* AS | 0.99 | 0.96-1.03 | 0.760 |
| *POR* MP | 0.96 | 0.92-1.00* | 0.030 |
| *SLCO1B1* AS | 1.01 | 0.99-1.03 | 0.064 |
| *SLCO1B1* MP | 1.02 | 1.00-1.03* | 0.032 |

**Figure 1.**
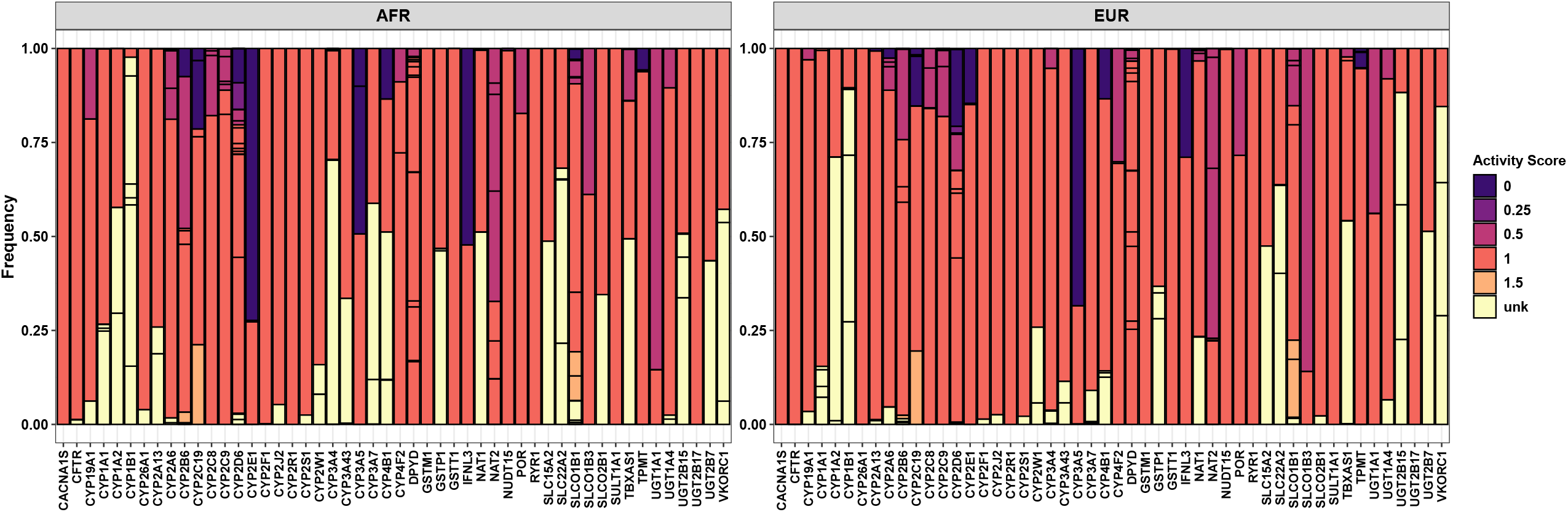
Star allele frequencies. Frequency of 262 * alleles observed in African (AFR) and European (EUR) ancestry participants of the UK Biobank (Table S4).

**Figure 2.**
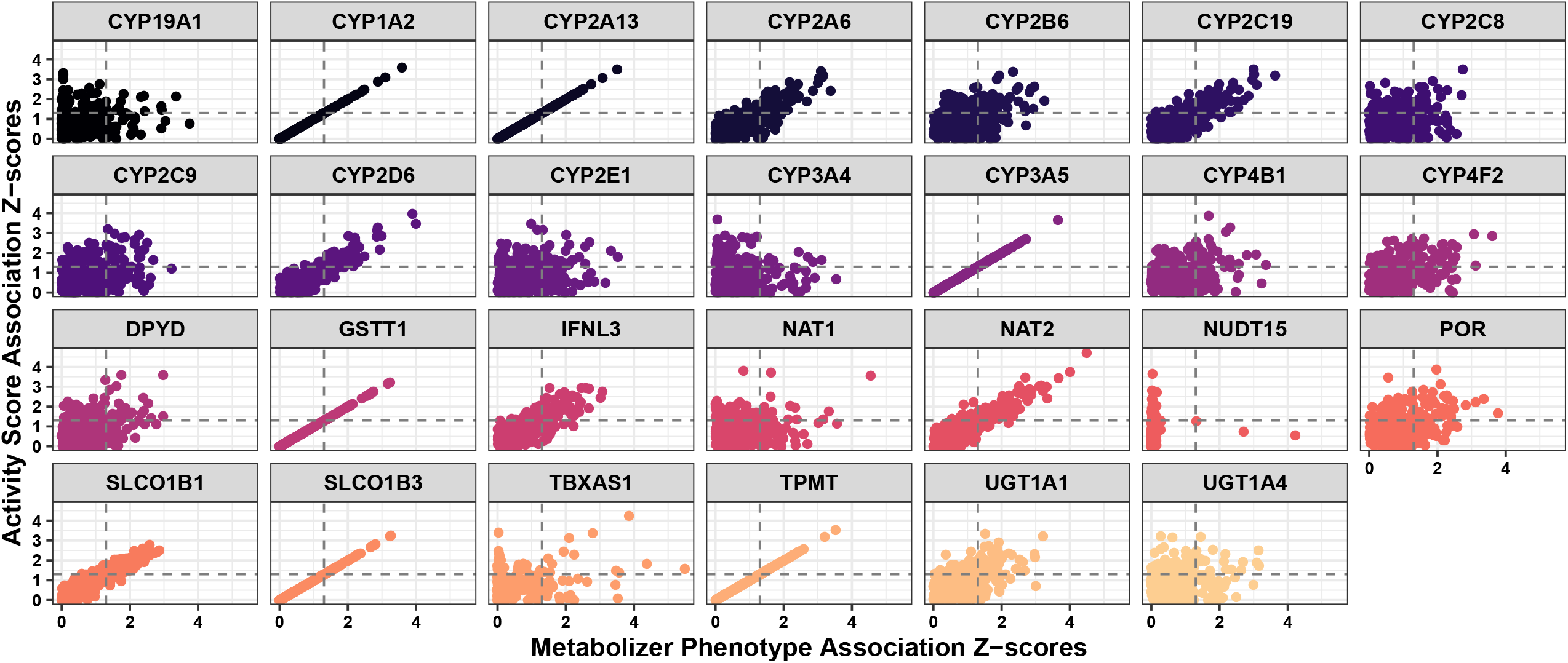
Metabolizer phenotype and activity score association tests. Association between activity score and metabolizer phenotype in UKB European ancestry participants. Each facet represents a single gene associated with 265 medication use phenotypes (data points); horizontal and vertical dashed lines indicate threshold p-value < 0.05. At six genes (*CYP1A2, CYP2A13, CYP3A5, GSTT1, SLCO1B3*, and *TPMT*), metabolizer phenotype and activity score capture the same level of variation making the correlation between x and y for these genes is 1

### *NAT2* haplotype dosage

Four common (frequency > 1%) *-alleles were detected at *NAT2* in EUR with *5 being the most frequent (45.2%) and the referent for all subsequent analyses even though *5 confers reduced activity to NAT2 protein product (Figure 3 and Table S4). The dose of each alternate haplotype (*1 (normal activity, 22.2% frequency), *6 (reduced activity, 29.5% frequency), and *7 (reduced activity, 2.31% frequency)) was regressed against simvastatin use covarying for sex, age, age^2^, sex*age, sex*age^2^, and 10 principal components of ancestry. Relative to *NAT2*\*5, dose of *NAT2*\*1 was significantly associated with increased odds of simvastatin use (*NAT2*\*1 OR=1.04, 95% CI=1.03-1.07, p=1.37×10^−5^; Figure 3). *NAT2*\*6 (OR=0.982, 95% CI: 0.961-1.00, p=0.109) and *7 (OR=0.941, 95% CI: 0.867-1.02, p=0.140) were not associated with simvastatin use.

**Figure 3.**
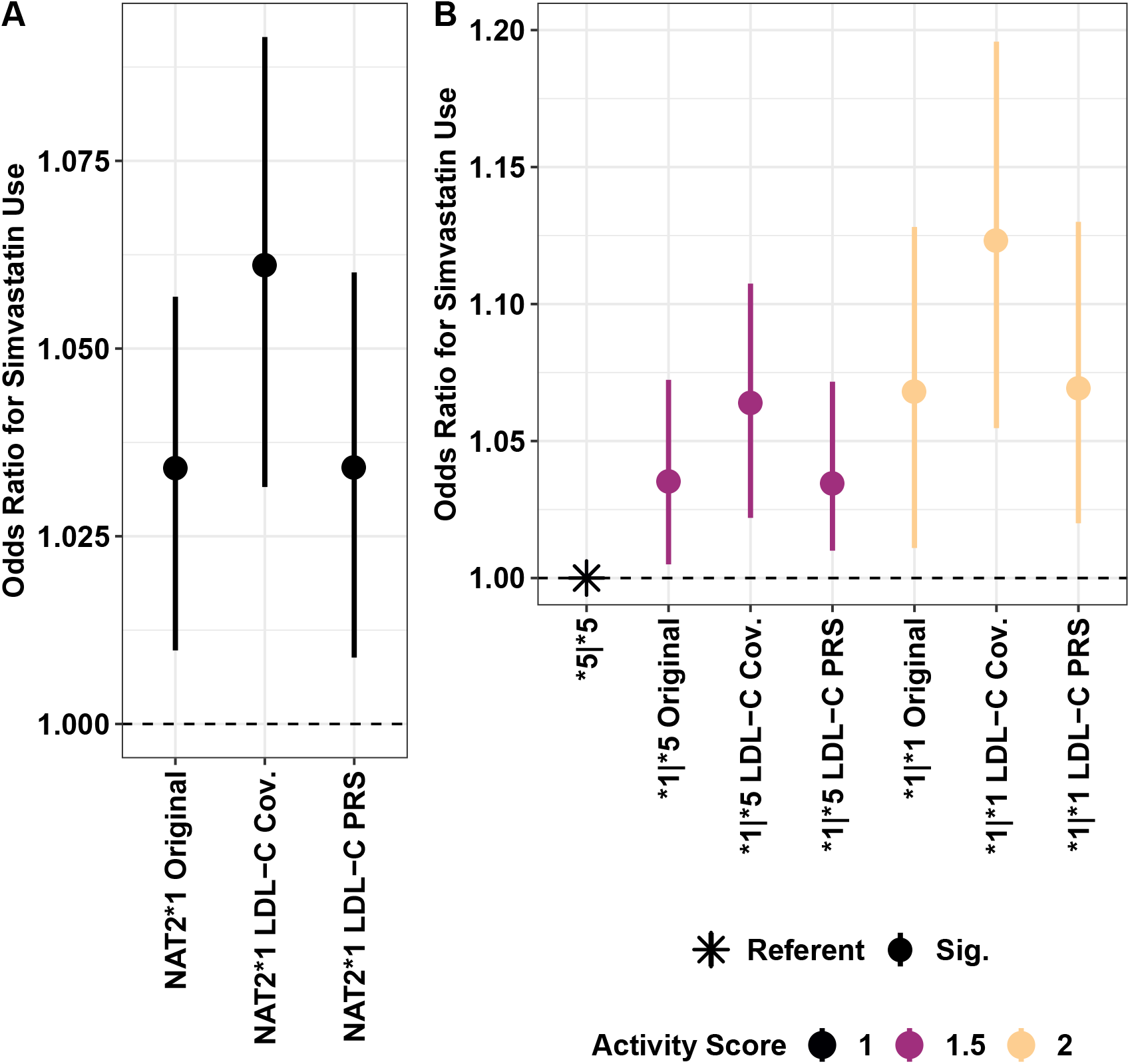
Main effect of *NAT2*\*1. (A) and diplotype (B) on simvastatin use. In both figures, the effect of NAT2 on simvastatin use was additional covaried with low-density lipoprotein cholesterol (LDL-C) levels (“LDL-C Cov.”) and polygenic risk scores for LDL-C levels (“LDL-C PRS”). Data points in (B) are colored by their activity score.

### *NAT2* and LDL-C

We next tested if the relationship between *NAT2* and simvastatin use was independent of conditions for which simvastatin is prescribed. *NAT2*\*1 dose remained associated with simvastatin use independently of LDL-C concentration and polygenic risk for LDL-C (Figure 4). The effect of *NAT2*\*1 on simvastatin use was independent of LDL-C (OR=1.07, 95% CI=1.05-1.09, p=1.14×10^− 8^) and PRS for LDL-C (*NAT2*\*1 given LDL-C PRS: OR=1.09, 95% CI=1.04-1.14, p=2.26×10^−4^). There was significant evidence of an interaction between *NAT2*\*1 dose and LDL-C concentration (*NAT2*\*1 dose OR=1.21, 95% CI: 1.06-1.38, p=0.005; *NAT2*\*1 dose × LDL-C OR: 0.957, 95% CI: 0.916-0.998, p=0.045; Figure 4). Among simvastatin users there was no difference in LDL-C concentration across *NAT2*\*1 diplotype (*NAT2*\*1|*1 = 2.74±0.652 mmol/L, *NAT2*\*1|*5 = 2.75±0.654 mmol/L, *NAT2*\*5|*5 = 2.75±0.671 mmol/L; ANOVA: F=0.386, df=2, p=0.680). In simvastatin non-users, the *NAT2*\*1|*5 and *1|*1 diplotypes were associated with increased LDL-C concentration relative to the *NAT2*\*5|*5 diplotype (*NAT2*\*1|*1 mean LDL-C concentration = 3.70±0.838 p=0.002, *NAT2*\*1|*5 = 3.68±0.838 p=2.10×10^−5^, *NAT2*\*5|*5 = 3.66±0.832; ANOVA: F=20.3, df=2, p=2.0×10^−9^).

**Figure 4.**
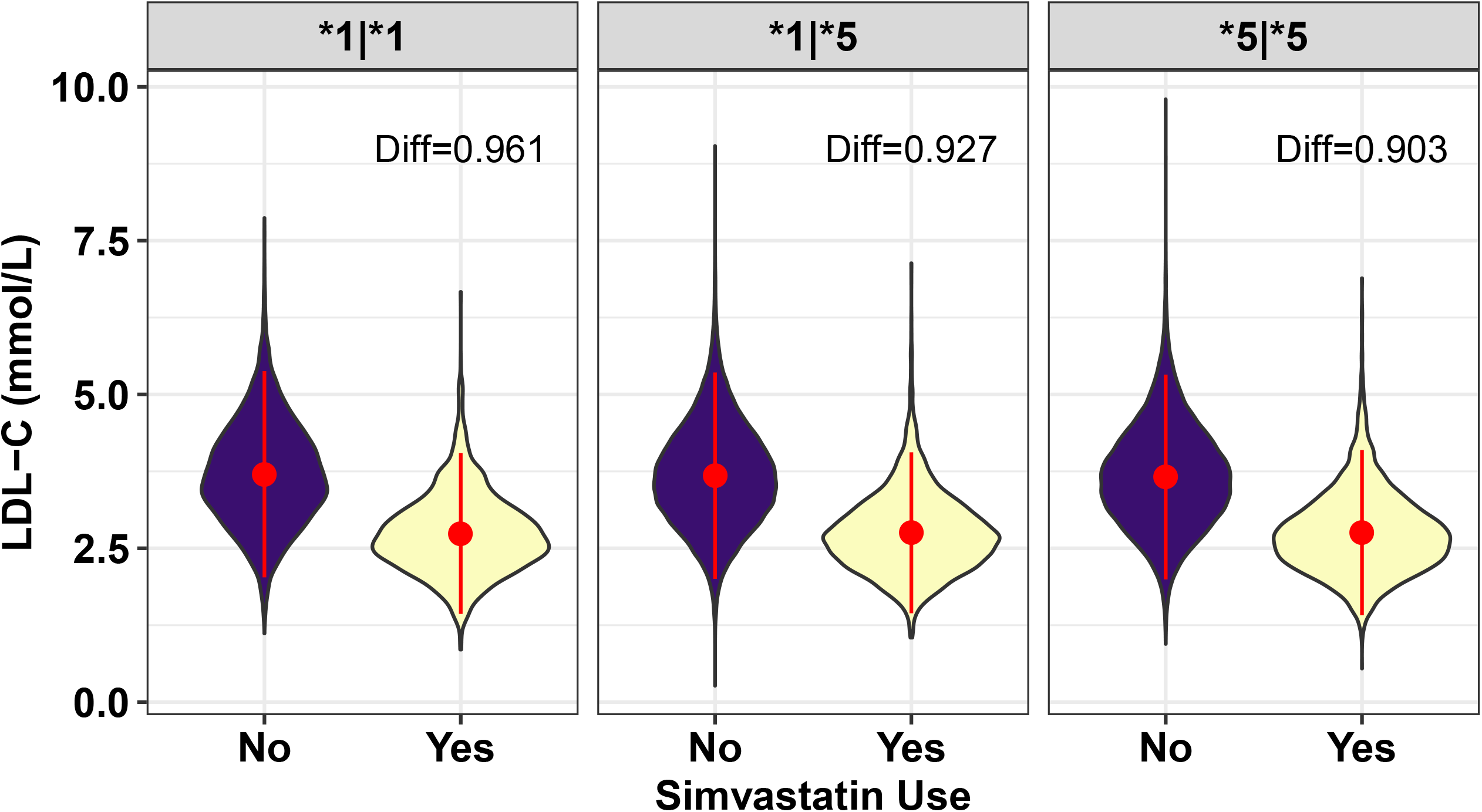
Interaction between *NAT2* and simvastatin use. Interaction between *NAT2*\*1 and simvastatin to lower concentration of low-density lipoprotein cholesterol (LDL-C; mmol/L). Each facet is labeled with the LDL-C concentration difference between simvastatin users and non-users. Data points and range represent mean and one standard deviation. “Diff” indicates the difference in LDL-C concentration between simvastatin users and non-users – all differences were significant (all comparisons were significant at p<0.05).

In an independent cohort of comparably aged participants (N=1,099 aged 40-70), we also observed that carriers of *NAT2*\*1 who use simvastatin show larger differences in LDL-C concentration compared to non-users (*NAT2*\*1 dose OR=1.09, 95% CI: 1.06-1.38, p=0.025; *NAT2*\*1 dose × LDL-C OR: 0.987, 95% CI: 0.976-0.998, p=0.029; Figure S4). Among individuals with *NAT2*\*5|*5, simvastatin users had mean LCL-C concentration 0.618 mmol/L lower than non-users (p=6.06×10^−6^); among individuals carrying a single copy of *NAT2*\*1, simvastatin users had mean LCL-C concentrations 0.679 mmol/L lower than non-users (p=6.03×10^−5^). The *NAT2*\*1|*1 condition was too rare to detect a statistically significant difference between LDL-C concentrations of simvastatin users (N=14) and non-users (N=133; p=0.064, LDL-C concentration difference=0.585 mmol/L).

In summary, as dose of *NAT2*\*1 increased, the difference in LDL-C between simvastatin users and non-users also increased (Figure 4). For *NAT2*\*1 carriers, this LDL-C concentration difference was more pronounced than that considering simvastatin alone: difference between users and non-users = 0.915 mmol/L.

Three additional statins were endorsed by UKB participants: atorvastatin (3.08% EUR case prevalence), rosuvastatin (0.63% EUR case prevalence), and pravastatin (0.48% EUR case prevalence). In **Supplementary Material** we describe the successful translation of findings from simvastatin use to the broad group of all statin users.

### Rare haplotype burden

*NAT2* diplotypes of known function were dummy coded as homozygous rare haplotype (N=17), heterozygous (N=4,575), and homozygous common haplotype (N=321,662). The dose of rare *NAT2* haplotypes of known function was not significantly associated with simvastatin use (OR=0.93, 95% CI: 0.84-1.02, p=0.145; see Figure S5 for results from 10,000 permutations).

### Cross-ancestry PGx effects on simvastatin use

*NAT2* AS (OR=0.969, 95% CI: 0.725-1.29, p=0.832) and MP (OR=0.890, 95% CI: 0.612-1.29, p=0.539) were not associated with simvastatin use in the UKB AFR cohort; however, three other pharmacogenes demonstrated nominally significant effects on simvastatin use (Table S8-S10): *CYP2A13, CYP2C8*, and *CYP4F2*. The dose of *CYP4F2*\*3 was associated with increased odds of simvastatin use relative to the *1|*1 reference diplotype (*1|*3 OR=1.32, 95% CI: 1.04-1.67, p=0.019 and *3|*3 OR=2.17, 95% CI: 1.15-3.91, p=0.013). However, this result was not independent of LDL-C concentration: *1|*3 OR=1.97, 95% CI: 0.673-5.89, p=0.219 and *3|*3 OR=0.151, 95% CI: 0.012-1.91, p=0.141.

## Discussion

Several outcomes are possible after taking a medication: (1) the user experiences the desired effects and no adverse events, (2) the user experiences the desired effect and adverse events, (3) the user experiences no therapeutic or adverse events, and (4) the user experiences no therapeutic effect but does experience adverse events. Understanding why these events occur and predicting which patients will experience no effect or adverse event scenarios is a critical barrier to personalized medicine complicated, in part, by the requirement for extremely large sample sizes in genetic studies of these complex traits related to substance and/or medication use and abuse.^5, 7, 23^ With the growing accessibility of curated large-scale biobanks and development of computational approaches to assign clinically actionable PGx variation^24-26^ this sample size barrier is now an advantage – large-scale biobanks exist in enormous sample sizes accompanied by deep phenotype data.^27, 28^ In this study, we extend previous reports of PGx variation at this scale^24, 25^ to understand how actionable variation at PGx loci associates with medication use. Using a gene-prioritization procedure, we identified a single association between *NAT2* and simvastatin use.

NAT2 is an acetyltransferase encoded by a highly polymorphic locus responsible for activation/deactivation of a large number of endogenous and exogenous compounds.^29, 30^ This variation causes individuals to be classified along a spectrum of slow to rapid metabolizers. The main effect detected was between carriers of the *NAT2* haplotypes *1 (defined by the absence of actionable variation in *NAT2*) and *5 (defined by the presence of rs1801280-C). The *NAT2* SNP rs1801280 has been associated with several adverse events due to its disruption of hydrophobic interactions among NAT2 side chains. Interestingly, it is known that SNPs associated with slower activity NAT2 confer different levels of slow activity inducing some heterogeneity among the slow-metabolizer population^31, 32^ that was not detected here – we observed no evidence of a relationship between the decreased activity haplotypes *NAT2*\*6 or *7 and simvastatin use relative to the *5 haplotype. This perhaps suggests that categorical MP information may be sufficient for translating these data clinically, although this hypothesis requires explicit testing.

Though implicated through candidate gene studies of LDL-C and detected via GWAS of total cholesterol,^18^ *NAT2* was only recently associated with LDL-C in a sample of ∼440,000 European individuals^19^ and the lead variant was associated with increased LDL-C concentration (β=0.017, p=4.60×10^−12^). *NAT2* also has been implicated in xenobiotic metabolism and skin fluorescence as a measure of advanced glycation end products.^33, 34^

We showed that the relationship between *NAT2* and simvastatin use survived the effects of LDL-C concentration and PRS for LDL-C. Furthermore, in combination with simvastatin use, *NAT2* diplotype was associated with ∼1-5% greater reduction in LDL-C concentration relative to simvastatin alone. Future work may consider gene-by-environment effects at *NAT2* risk loci to identify relationships between simvastatin use, *NAT2*, and modifiable non-pharmacological risk factors (e.g., exercise, diet, life experiences)^35, 36^ that may modify the effect of *NAT2* and highlight more effective strategies for reducing LDL-C concentration and/or mitigating the effects of polygenic risk for LDL-C concentration.^37, 38^ In combination, genetic and environmental information regarding simvastatin efficacy may maximize desirable outcomes (outcome 1 described above) while minimizing potentially adverse or undesirable outcomes (outcomes 2-4 described above).

Based on our gene-prioritization procedure, *NAT2*\*1-simvastatin use was the only observed association after multiple testing correction. We notably did detect several additional gene-medication use associations involving either MP or AS but not both. Though AS and MP capture similar degrees of phenotype variation, we focused our analysis on deeply characterizing only the most reliable association. This lack of support for PGx-based association with medication use phenotypes perhaps stems from the UK Biobank being a generally healthy cohort and, by extension, lacking enrichment for deleterious PGx variation.

Several additional PGx loci exhibited suggestive evidence of an effect on simvastatin use, some of which have prior empirical support: *CYP2C8, CYP2C9*, and *CYP3A5* are directly involved in simvastatin to metabolite conversion,^39^ and *SLCO1B1* was associated with increased risk of statin induced myopathy.^40^ Though *CYP2D6* has not been directly linked to simvastatin response, *POR* (P450 oxidoreductase) has been previously associated with atorvastatin metabolite production. It is therefore unsurprising that several cytochrome P450 enzymes show an effect on statin metabolism. There is no study, to our knowledge, supporting the clinical utility of a polygene prediction of simvastatin use or efficacy; however, the findings from this study, and those from polygenic risk score studies, suggest that this approach may be feasible and possibly more informative than single gene prediction of medication response outcomes.^4, 41^ Furthermore, there is currently no recommendation for simvastatin dose using phase II metabolism enzymes (https://go.drugbank.com/drugs/DB00641) – we provide support for investigating the combined effects of phase I and phase II metabolism enzymes on simvastatin metabolism.

This study has three primary limitations. First, the UK Biobank is considered a generally healthy population-based cohort enriched for participants older than the general population. To mitigate these effects we covaried with several measures of age (i.e., age, age^2^, sex×age, sex×age^2^). Second, in this study we do not consider the effects of polypharmacy (e.g., number of medications used^2^). Indeed, many UKB participants endorse use of more than one medication and addressing this observation is an essential research and clinical question but is a tricky topic to address. Future work will require careful consideration of variable medication prescription frequencies across regions and patient care networks (e.g., combined statin and antidepressant perscription^42^), generalizability outside the UK (i.e., prescription combinations may differ by country), and how to classify medications containing more than one active compound (e.g., combined opioid and acetaminophen analgesics). Third, PGx loci are occasionally duplicated/deleted and the frequency of these events various by population and genomic region. In favor of increased sample size, we focused our analysis on PGx variation called directly from SNP-array data;^10, 11, 24-26^; however, some diplotypes, ASs, and MPs may change if sequence-based *-allele assignments are made. As larger sample sizes of whole-genome and/or whole-exome sequencing data become available, the power of PGx-wide association testing will drastically improve.

## Summary/Conclusions

We investigated pharmacogenetic correlates of medication use phenotypes in the UK Biobank. Though a generally healthy cohort, we identified hundreds of clinically actionable PGx variants consistent with previous investigations.^24, 25^ By characterizing the relationship between *NAT2* and simvastatin (and other statin) use, we uncover an interactive relationship between the two resulting in larger reduction in LDL-C concentration. This finding, among dozens of analysis-wide PGx-medication associations, highlights the utility of large-scale biobanks for understanding and identifying genetic variation with evidence for clinically actionable prescription recommendations.

## Data Availability

All data referred to in this manuscript are provided as supplementary information. Individual level data may be accessed by bone fide researchers through the UK Biobank. This research has been conducted using the UK Biobank Resource (application reference no. 58146).

https://www.ukbiobank.ac.uk/data-showcase/

https://www.ncbi.nlm.nih.gov/projects/gap/cgi-bin/study.cgi?study_id=phs000906.v1.p1

## Abbreviations

AFR: African ancestry
AS: activity score
EUR: European ancestry
GWAS: genome-wide association study
LDL-C: low-density lipoprotein cholesterol
MP: metabolizer phenotype
PGx: Pharmacogene
PRS: polygenic risk score
UKB: United Kingdom Biobank
VCF: variant call format (file type)
VIP: very important pharmacogene

## Acknowledgements

This research has been conducted using the UK Biobank Resource (application reference no. 58146).

## Sources of funding

This study was supported by the National Institutes of Health (R21 DC018098 (RP), R21 DA047527 (RP), and F32 MH122058 (FRW)).

## Disclosures

EJM reports grants from Bracco and Eidos, consulting for General Electric, Alnylam, and Pfizer. DJ has served as consultant and steering committee member for MyoKardia, Inc.

## Supplementary Materials

### Supplementary Results

#### Analysis-wide associations in European ancestry

The *NAT2*-simvastatin use relationship was the only phenotype surviving multiple testing correction in activity score and metabolizer phenotype association tests. However, among UKB EUR participants, four gene-medication use associations survived analysis-wide multiple testing correction (i.e., they were significant after multiple testing correction in metabolizer phenotype associations but not in activity score associations; Table S6). These associations were between (1) *TBXAS1* and telmisartan use (MP OR=0.058, 95% CI: 0.024-0.192, p=3.48×10^−8^ and AS OR=0.692, 95% CI: 0.428-1.23, p=0.170), (2) *TBXAS1* and Pulmicort inhaler use (MP OR=0.077, 95% CI: 0.029-0.314, p=1.29×10^−5^ and AS OR=0.622, 95% CI: 0.395-1.06, p=0.058), (3) *NAT1* and rabeprazole sodium use (MP OR=0.073, 95% CI: 0.027-0.298, p=9.27×10^−6^ and AS OR=0.681, 95% CI: 0.461-1.06, p=0.071), and (4) *NUDT15* and iron product use (MP OR=0.004, 95% CI: 3.47×10^−4^-0.100, p=2.40×10^−4^ and AS OR=1.54, 95% CI: 0.428-9.86, p=0.579).

We next investigated common pharmacogene variation at these loci and uncovered several effects of minor haplotype dose on medication use: *TBXAS1* and telmisartan use dose OR=1.44, 95% CI: 0.813-2.33, p=0.170; *1|*8 OR=0.653, 95% CI: 0.257-2.33, p=0.304; *8|*8 OR=16.9, 95% CI: 5.11-40.9, p=4.18×10^−8^; *TBXAS1* and Pulmicort inhaler use dose OR=1.61, 95% CI: 0.942-2.53, p=0.058; *1|*8 OR=1.07, 95% CI: 0.530-1.92, p=0.832; *8|*8 OR=13.1, 95% CI: 3.20-34.9, p=1.27×10^−5^; and *NAT1* and rabeprazole sodium use dose OR=1.47, 95% CI: 0.941-2.17, p=0.071; *1|*14 OR=1.12, 95% CI: 0.661-1.77, p=0.646; *14|*14 OR=13.8, 95% CI: 3.37-37.2, p=8.89×10^−6^. Note that in European ancestry participants, *NUDT15* * alleles were too rare to test for association with iron product use (Table S4).

Pharmacogene dose effects on medication use meeting nominal significance (Table S10) highlight eight medications (antifungals, antihypertensives, and benzodiazepines) with suggestive evidence for effect of pharmacogenetic variation.

#### Simvastatin Effects Translated to All Statins

We next considered whether the effects of *NAT2* on simvastatin translated to the more general group of statin users. We incorporated endorsement of three additional statin the analysis: atorvastatin (3.08% EUR case prevalence; MP OR=1.06, 95% CI: 1.02-1.11, p=0.005 and AS OR=1.10, 95% CI: 1.03-1.18, p=0.004), rosuvastatin (0.63% EUR case prevalence; MP OR=1.02, 95% CI: 0.923-1.11, p=0.745 and AS OR=1.03, 95% CI: 0.890-1.20, p=0.655), and pravastatin (0.48% EUR case prevalence; MP OR=1.03, 95% CI: 0.936-1.15, p=0.486 and AS OR=1.08, 95% CI: 0.914-1.28, p=0.359).

*NAT2* AS and MP were significantly associated with any statin use (MP OR=1.06, 95% CI: 1.04-1.08, p=2.65×10^−8^ and AS OR=1.11, 95% CI: 1.07-1.14, p=4.13×10^−9^). Relative to *NAT2*\*5, the dose of *NAT2*\*1 was associated with increased odds of statin use (OR=1.11, 95% CI: 1.07-1.14, p=0.001) independently of LDL-C (*NAT2*\*1 OR=1.27, 95% CI: 1.11-1.45, p=3.62×10^−4^) and polygenic risk for LDL-C (*NAT2*\*1 OR=1.04, 95% CI: 1.01-1.06, p=0.001). We detected an interaction between *NAT2*\*1 and LDL-C (*NAT2*\*1 dose × LDL-C OR=0.946, 95% CI: 0.907-0.987, p=0.011) but not polygenic risk for LDL-C (*NAT2*\*1 dose × LDL-C OR=816.0, 95% CI: 0.15-4.03×10^7^, p=0.396).

#### Medication use in African ancestry participants

In the UKB African ancestry sample (N=7,649), we detected 13 nominally significant gene-medication use combinations (p < 0.05; Table S7). Four of these involved *CYP2A13*, at which we did not observe common haplotypes of known function and were therefore excluded from dose association. Using nominally significant findings from the AFR population, we identified putative effects of *CYP2B6* on ramipril use, *CYP4B1* on use of aspirin, omeprazole, and lansoprazole use, *CYP4F2* on simvastatin use, and *UGT1A1* on metformin and aspirin use (Table S8-S10).

### Supplementary Figures

**Figure S1.**
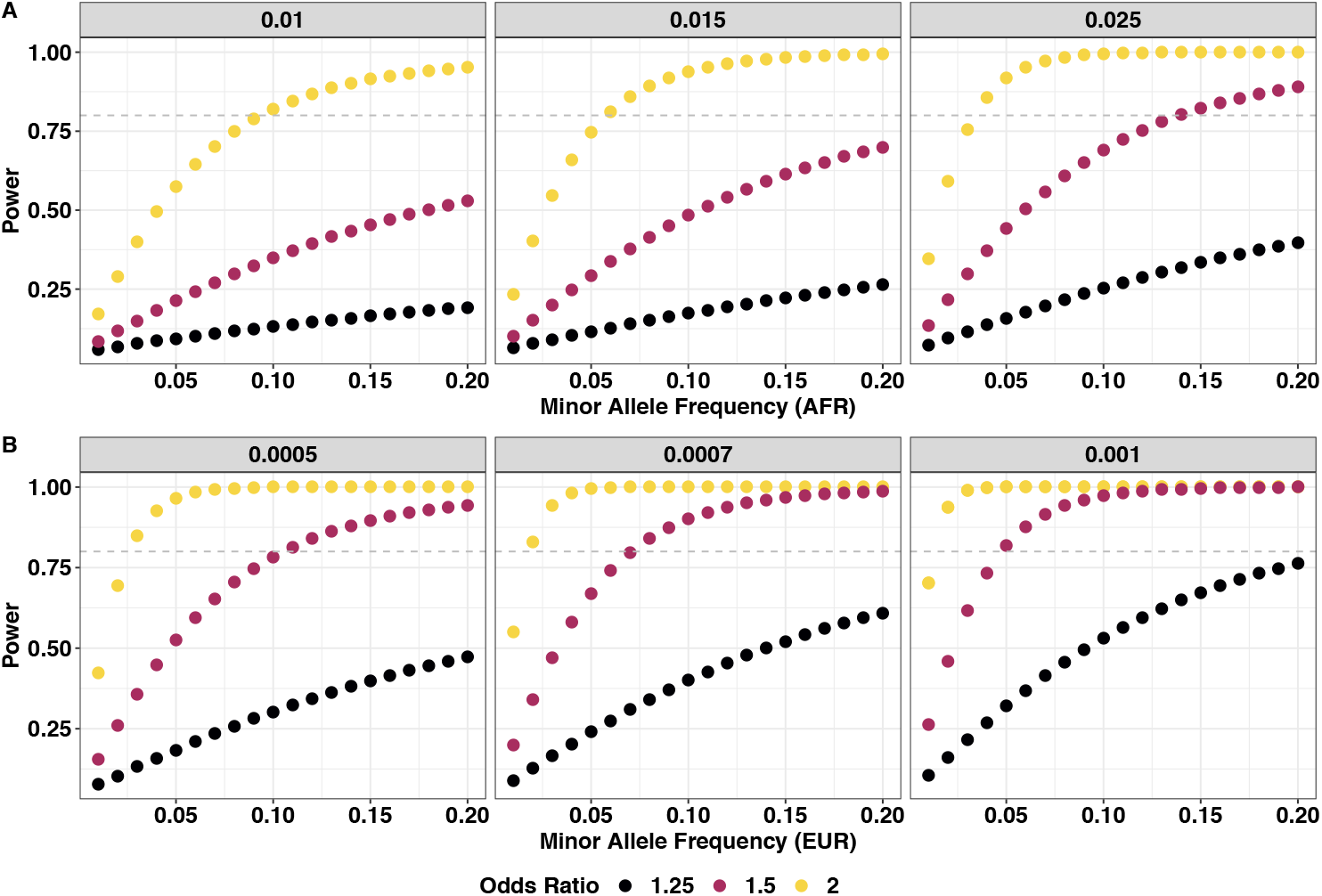
Power analysis in AFR (A) and EUR (B) at medication use case prevalence values indicated in each facet label. Grey dashed lines indicate 80% power.

**Figure S2.**
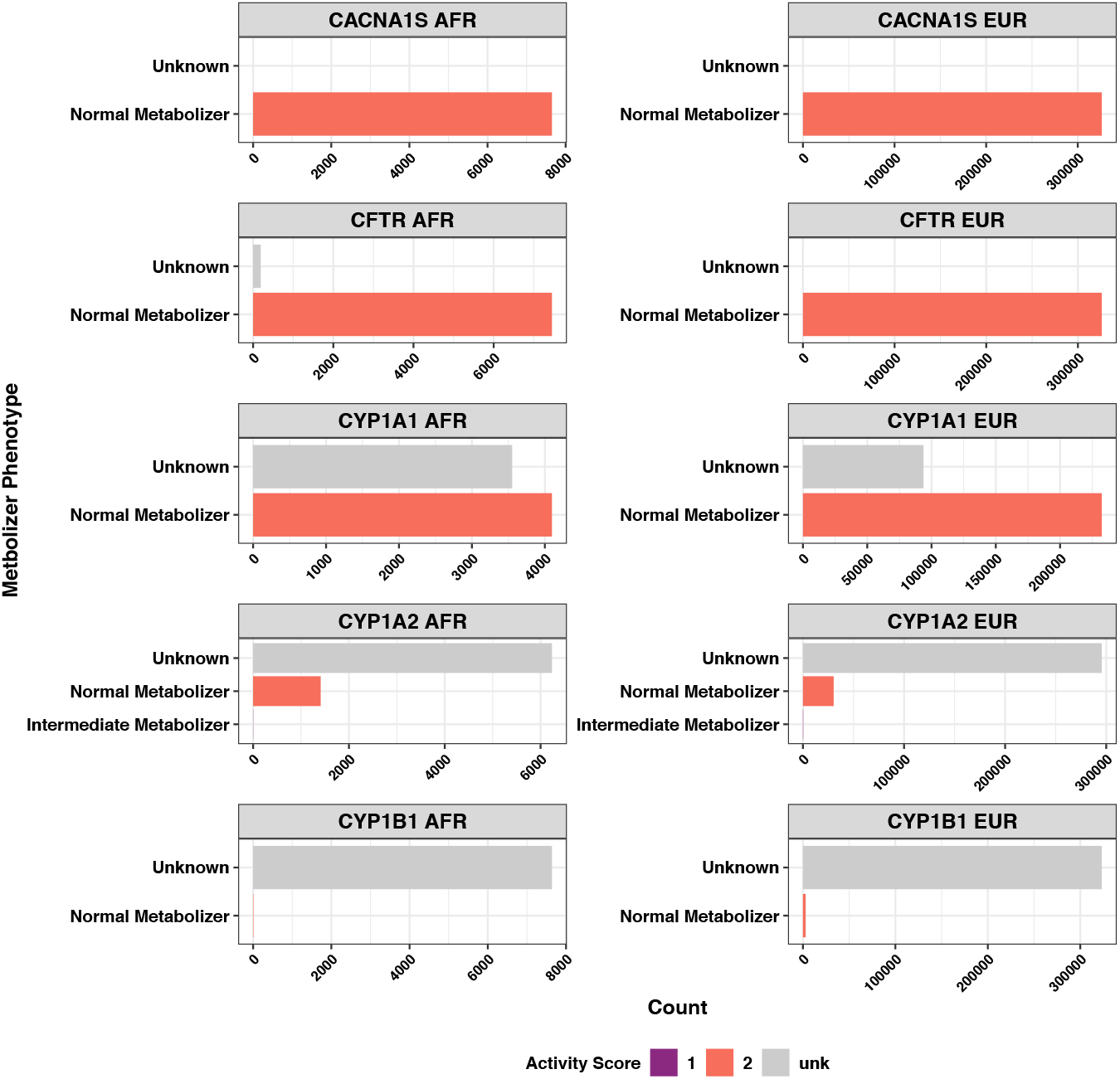
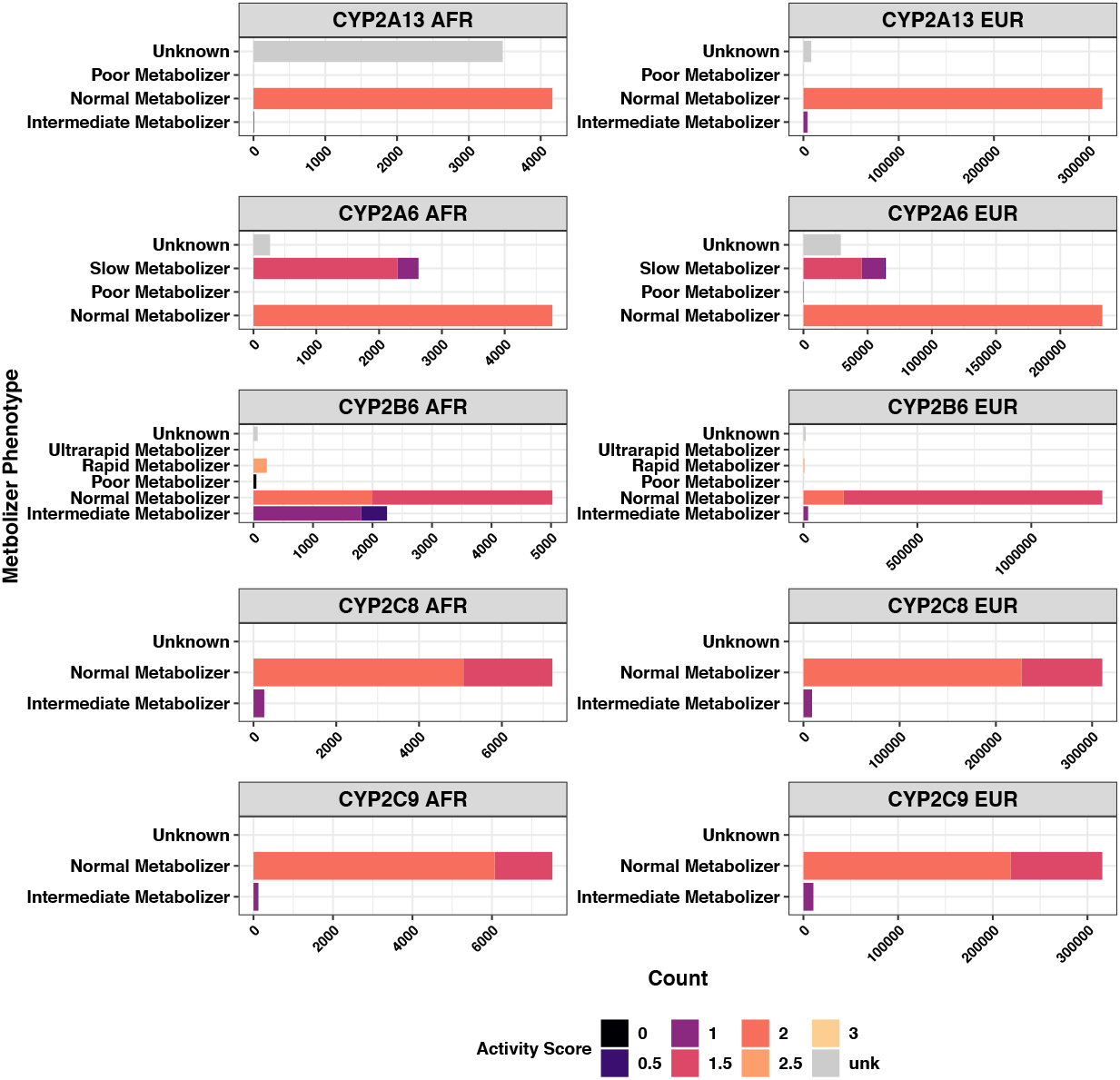
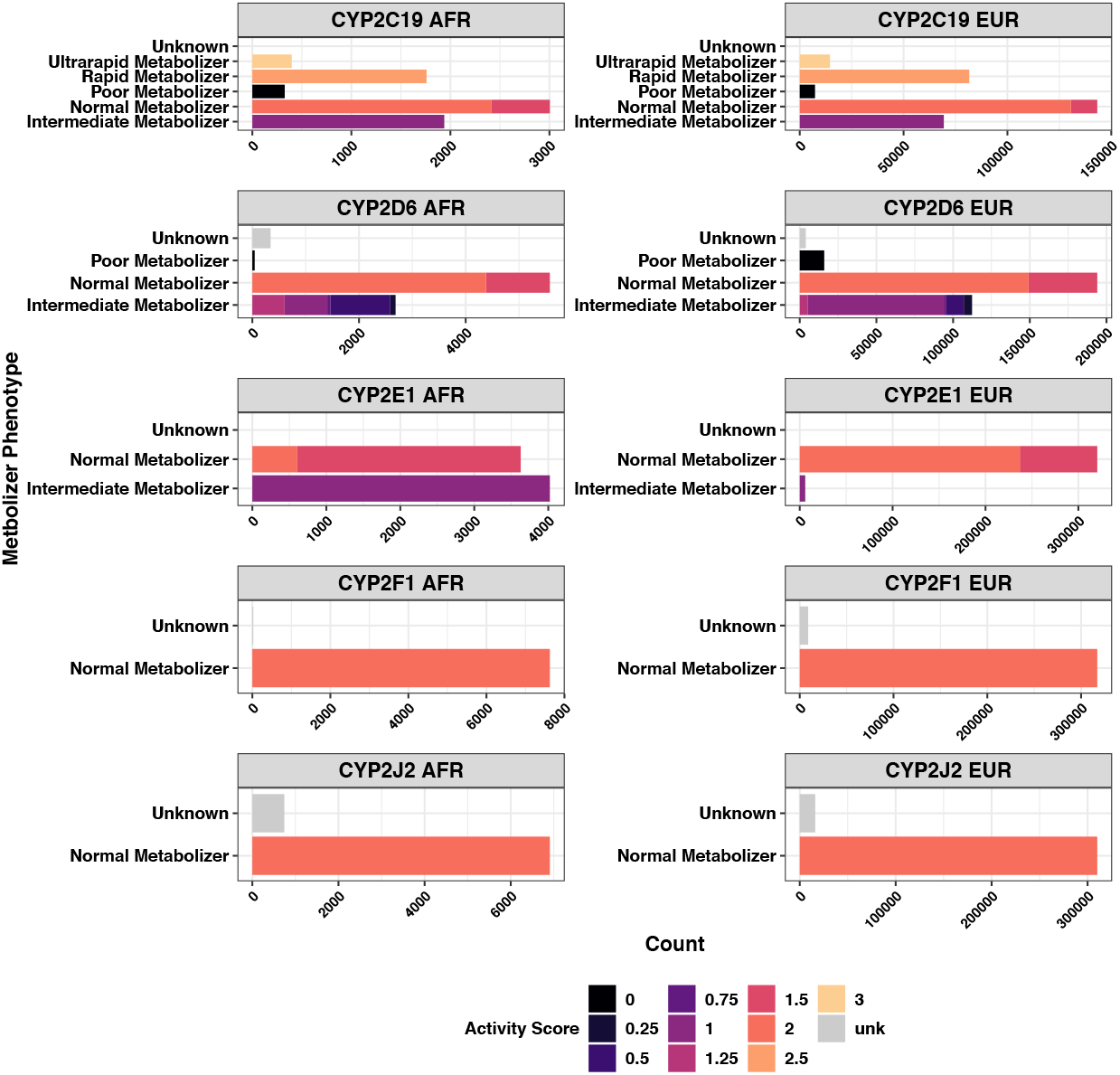
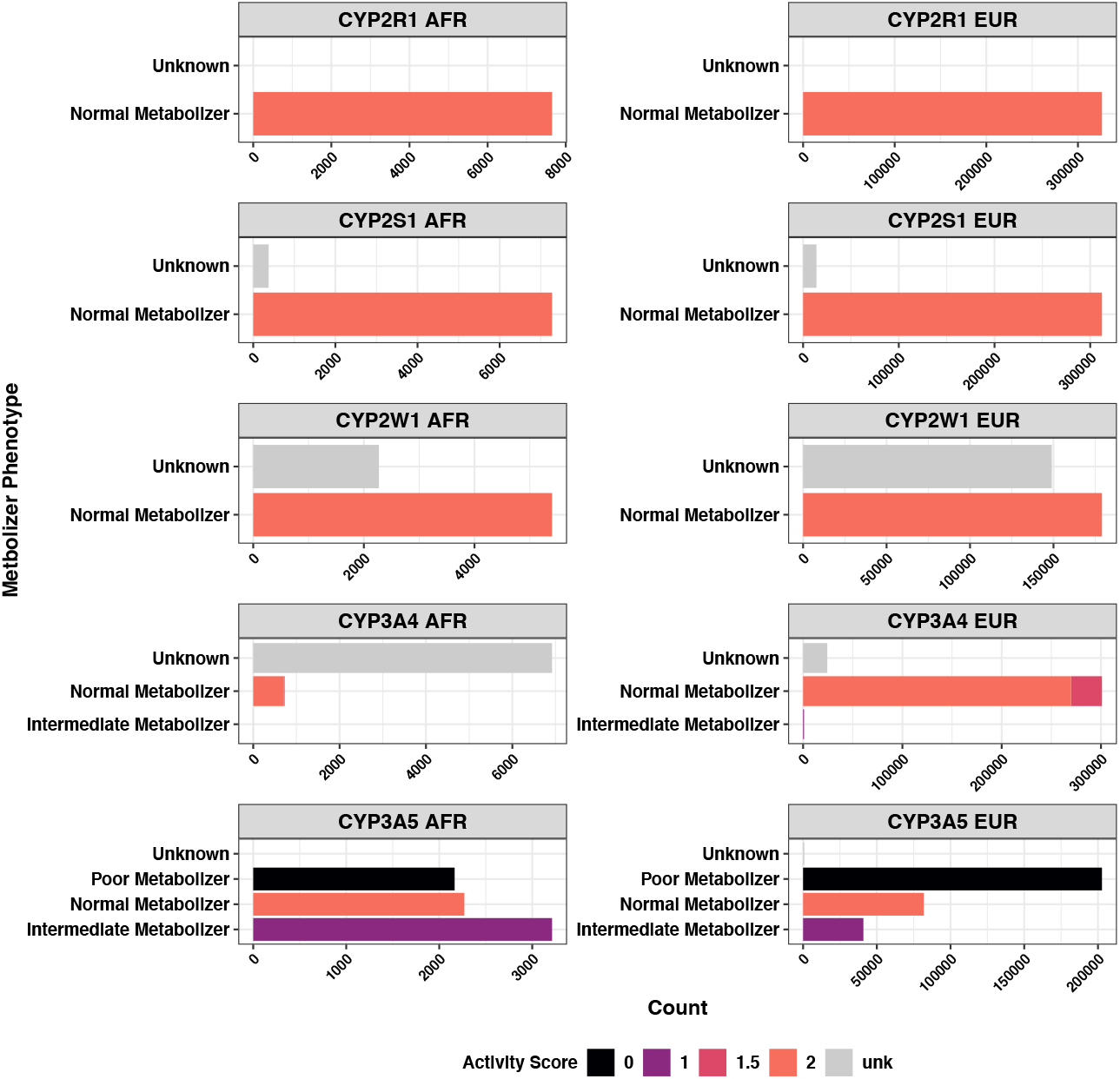
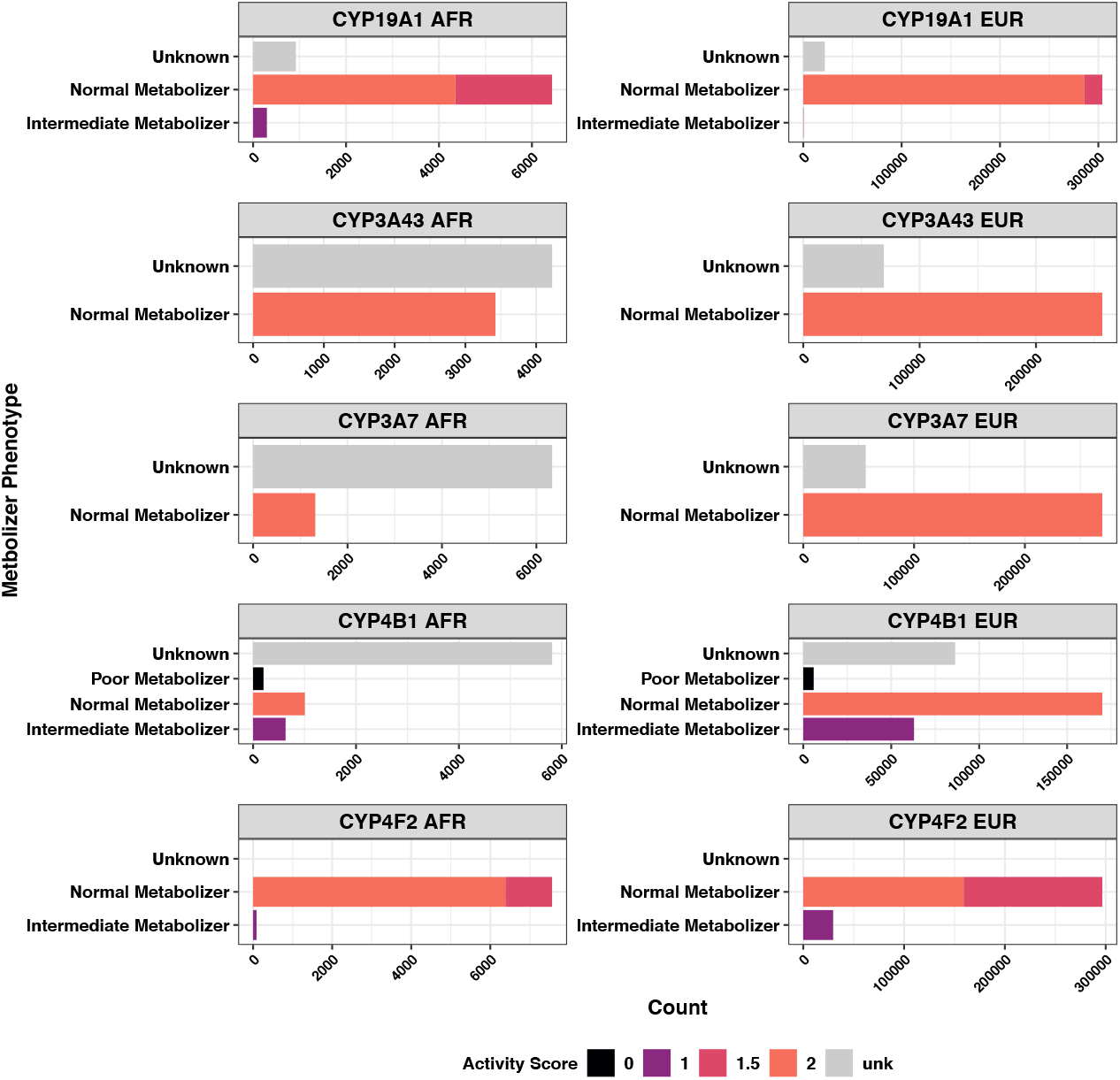
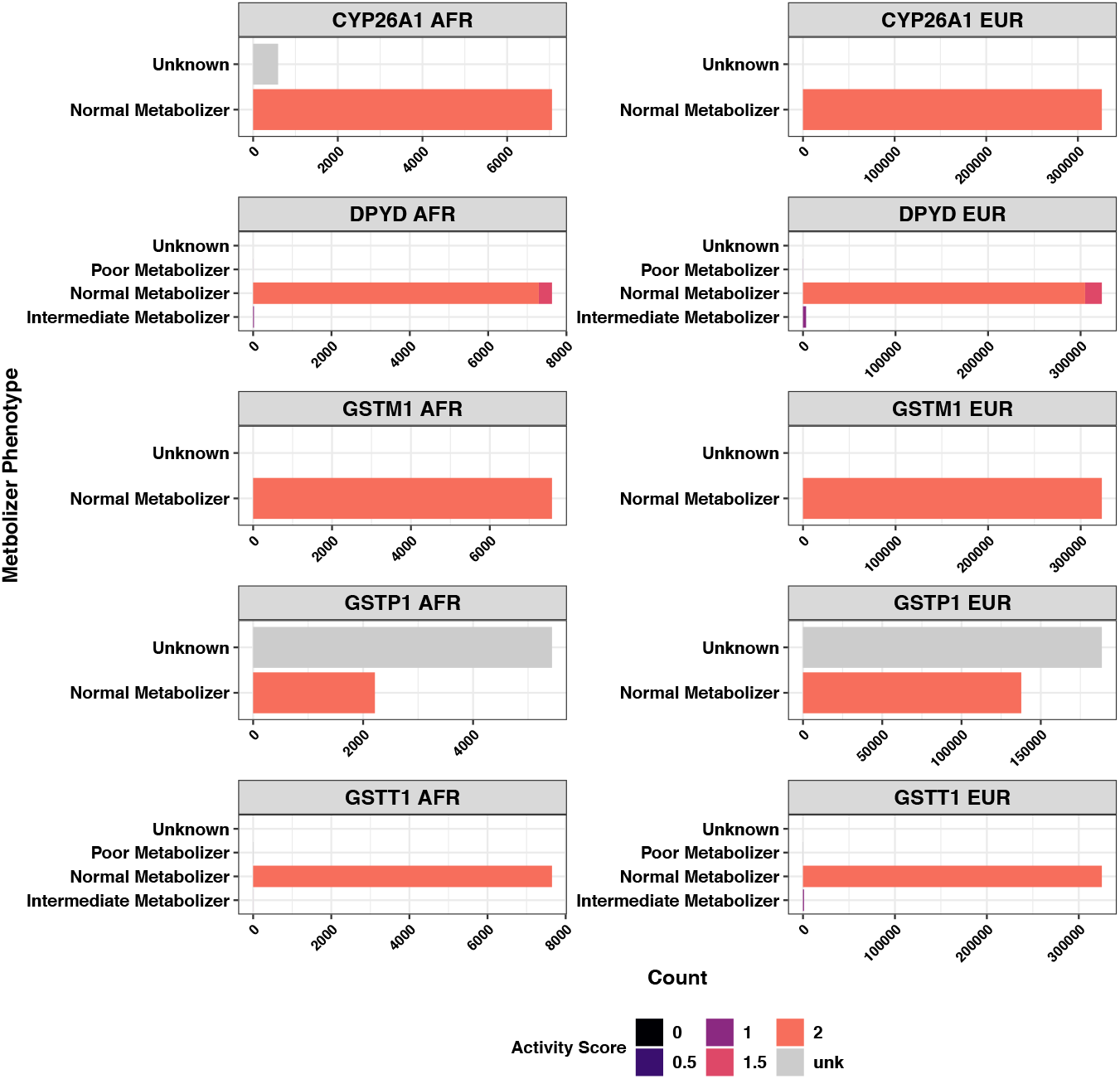
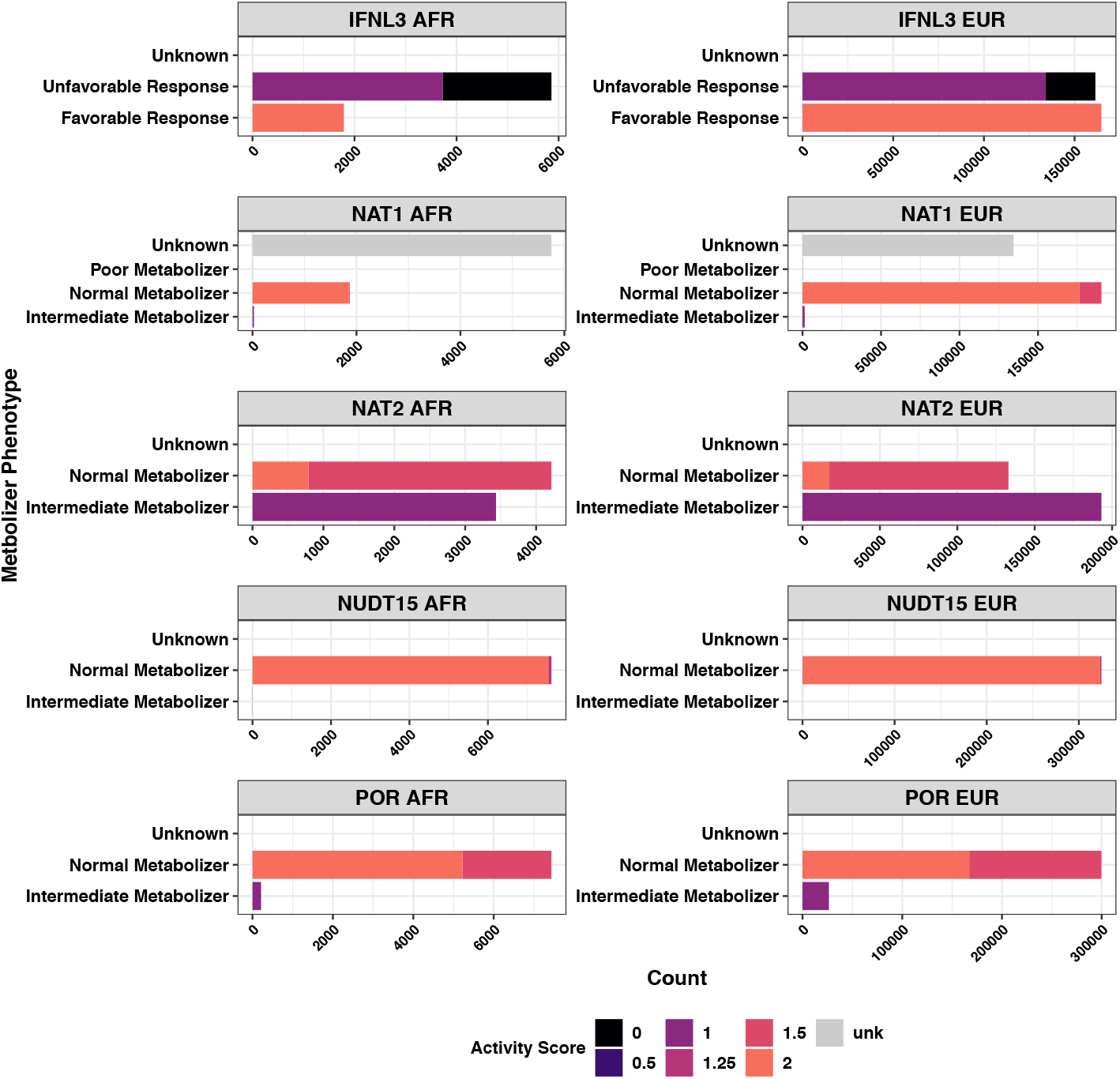
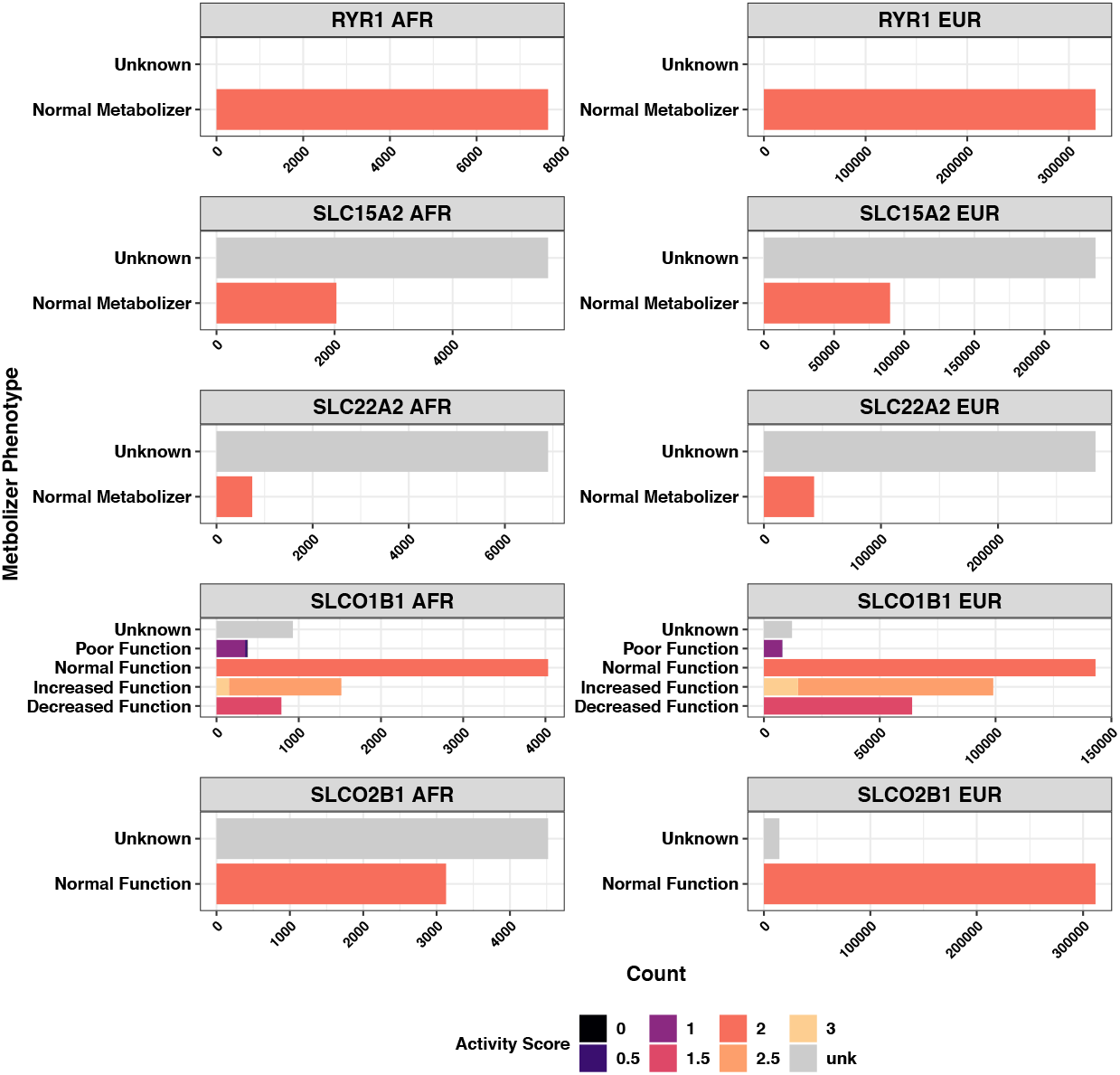
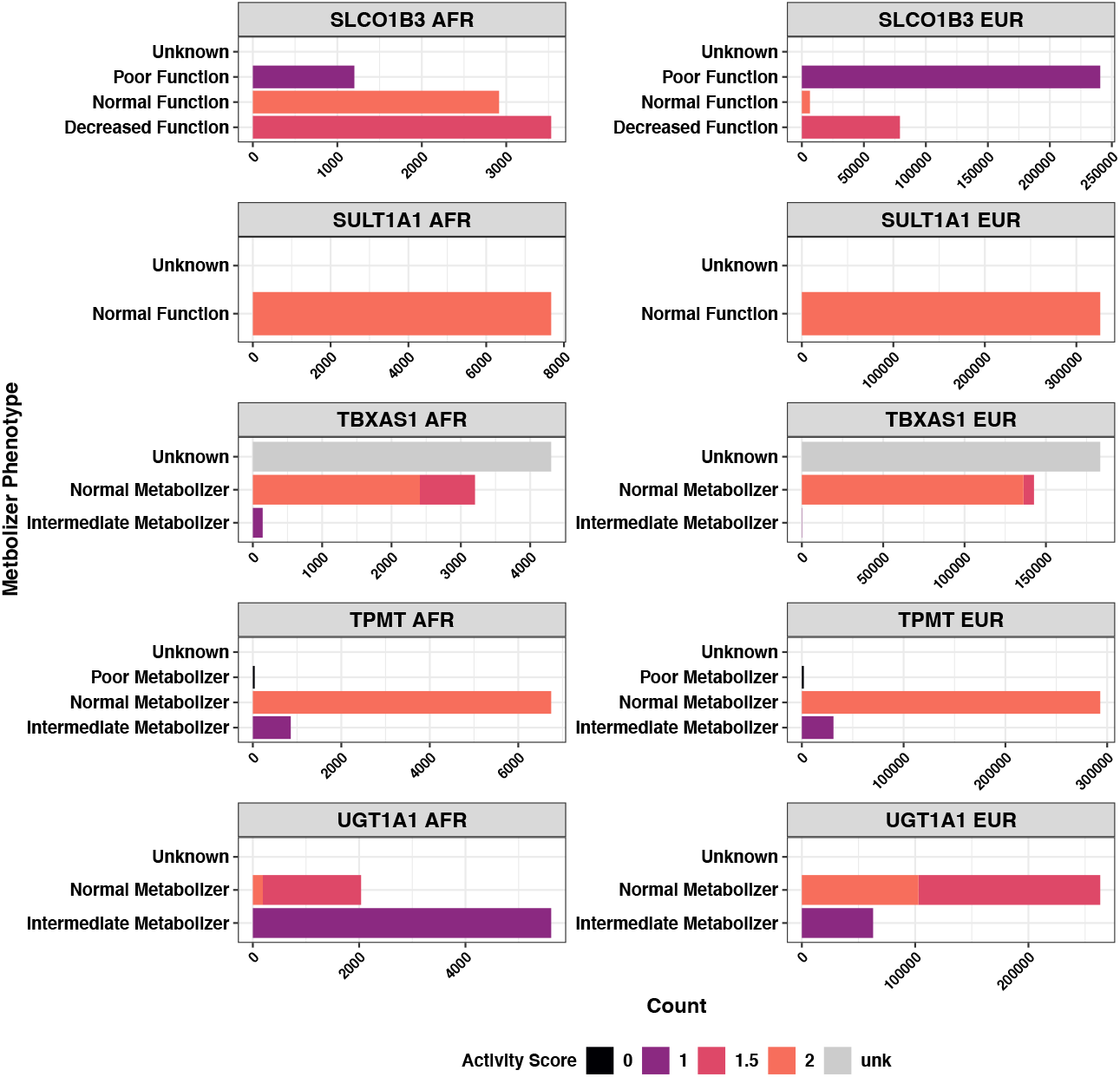
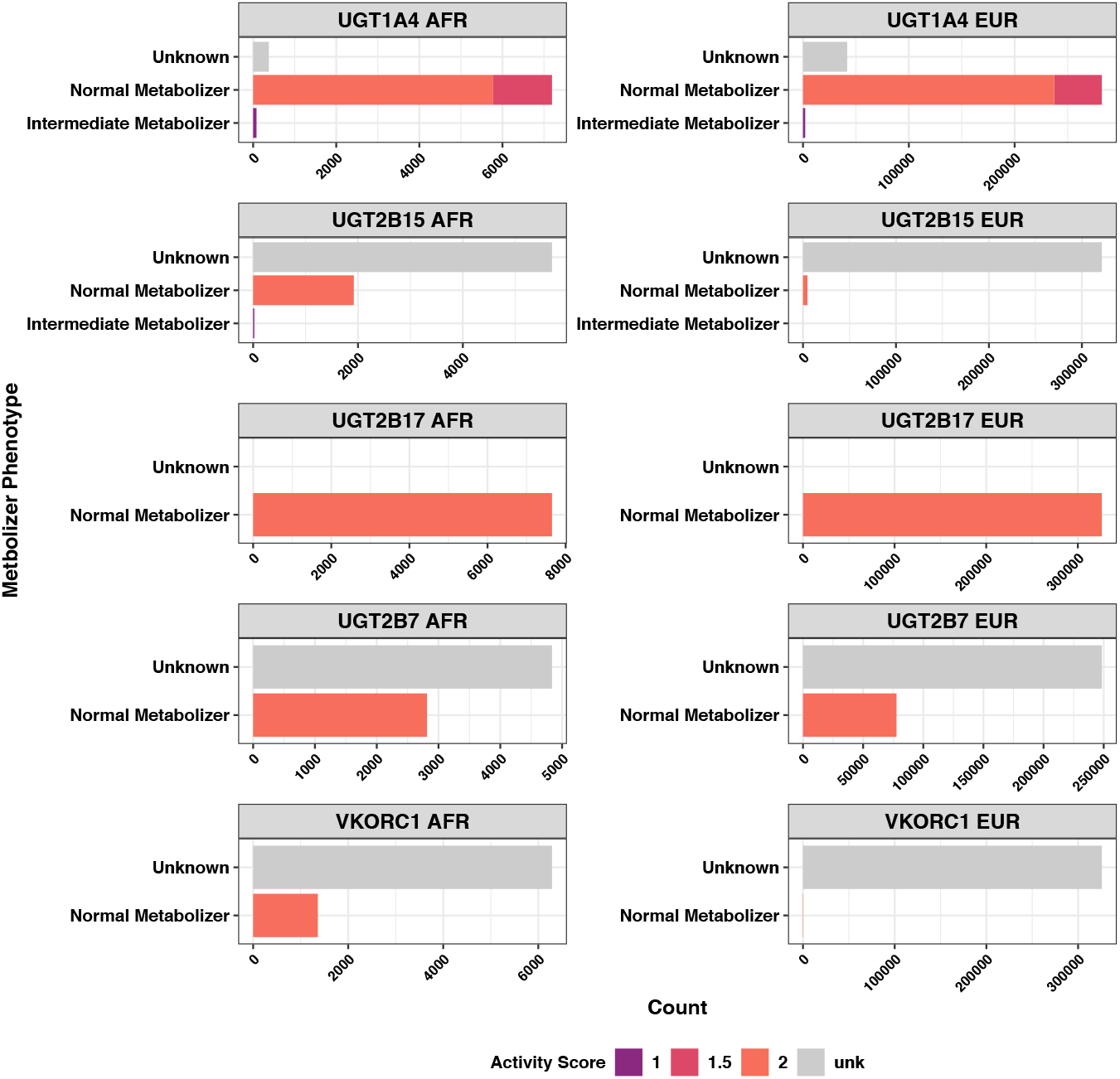
Pharmacogene phenotype distribution in African (AFR, left column; N = 7,649) and European (EUR, right column, N = 326,214) ancestry participants of the UK Biobank.

**Figure S3.**
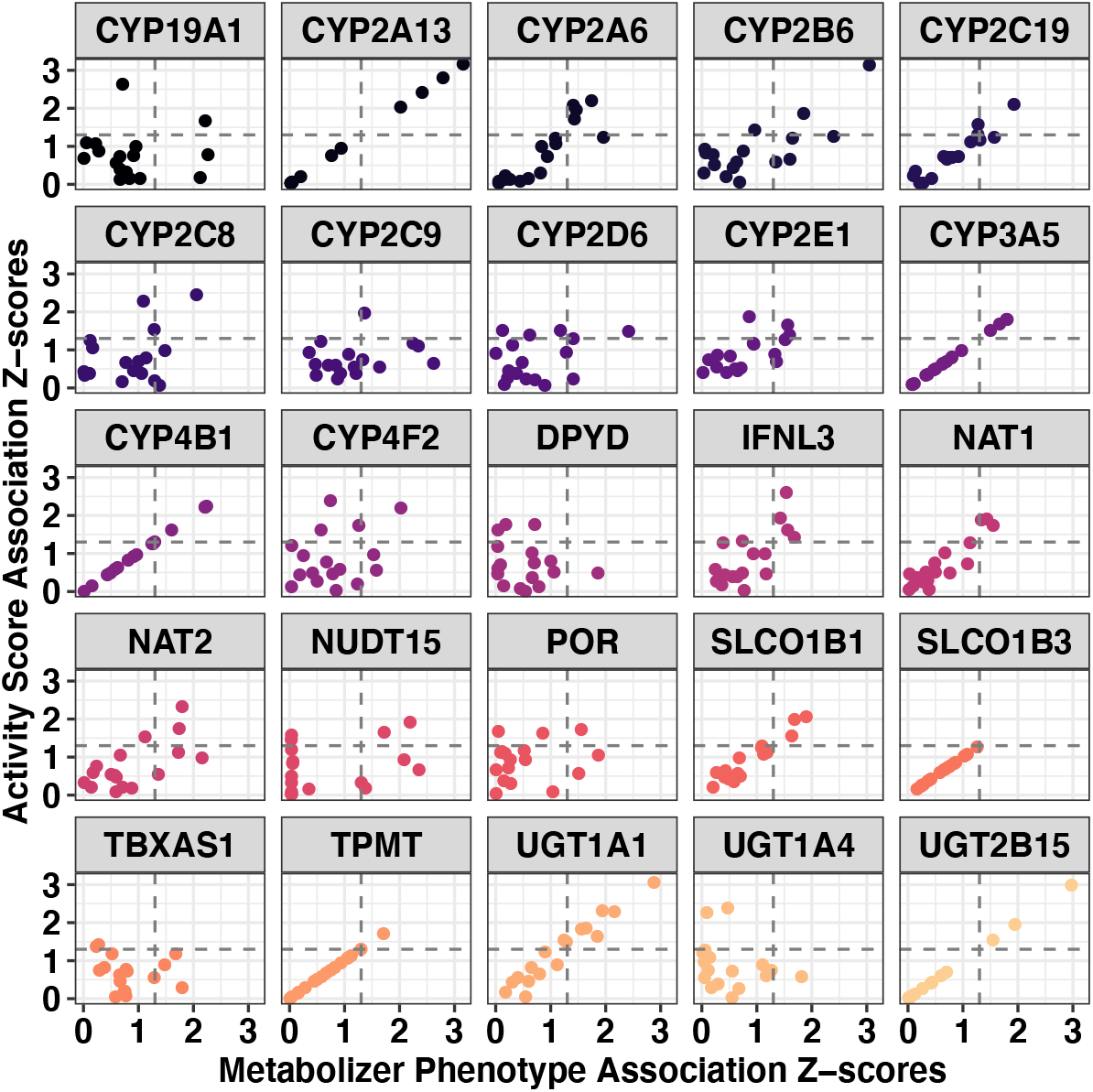
Association between activity score and metabolizer phenotype in UKB African ancestry participants. Each facet represents a single gene associated with 326 medication use phenotypes (data points); horizontal and vertical dashed lines indicate threshold p-value < 0.05. At six genes (*CYP2A13, CYP3A5, CYP4B1, SLCO1B3, TPMT*, and *UGT2B15*), metabolizer phenotype and activity score capture the same level of variation making the correlation between x and y for these genes is 1.

**Figure S4.**
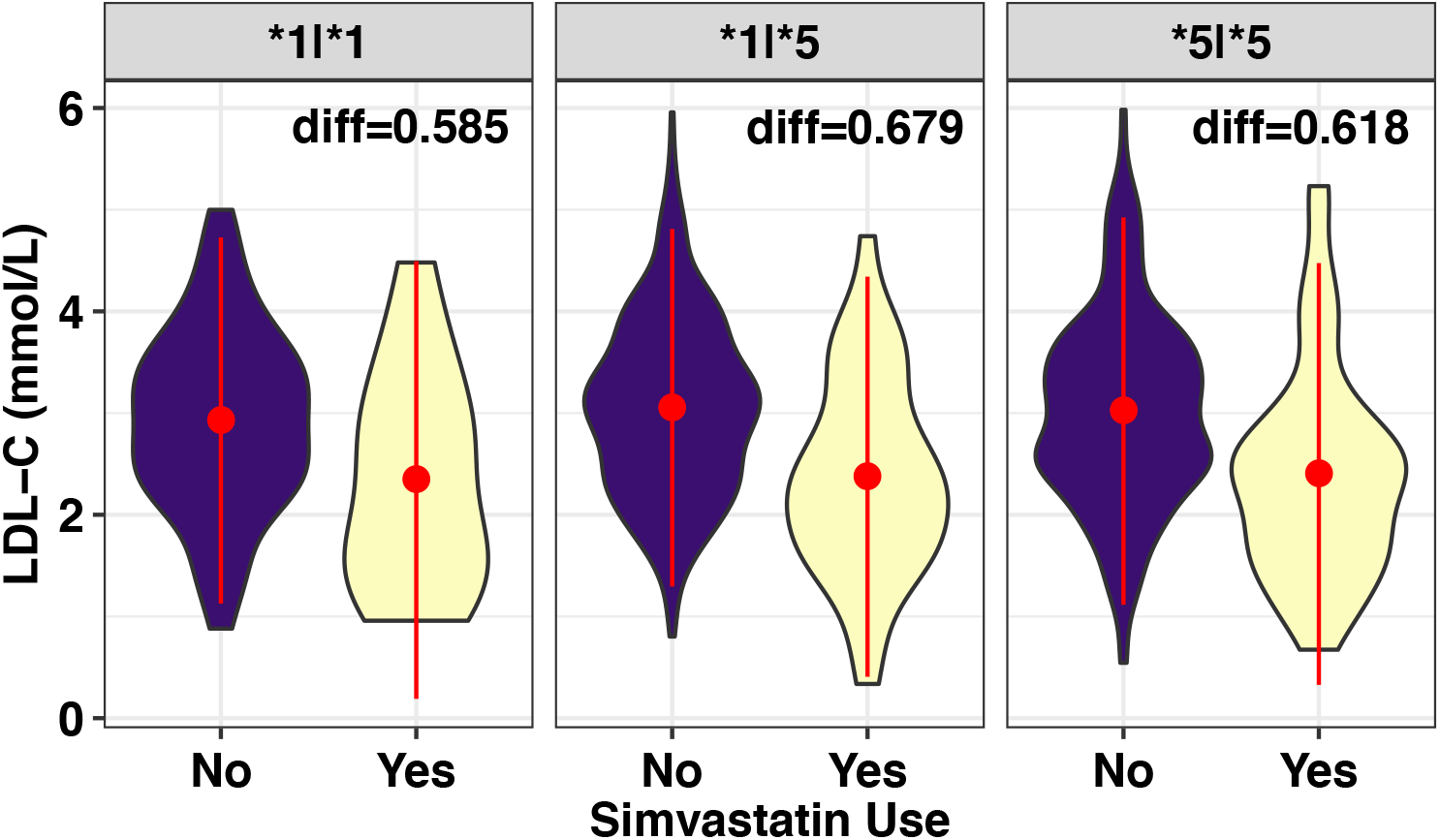
Interaction between *NAT2*\*1 and simvastatin to lower concentration of low-density lipoprotein cholesterol (LDL-C; mmol/L) in 1,099 individuals from the eMERGE Pharmacogenomics Pilot. Each facet is labeled with the LDL-C concentration difference between simvastatin users and non-users. Data points and range represent mean and one standard deviation. “Diff” indicates the difference in LDL-C concentration between simvastatin users and non-users – all differences were significant (all comparisons were significant at p<0.05).

**Figure S5.**
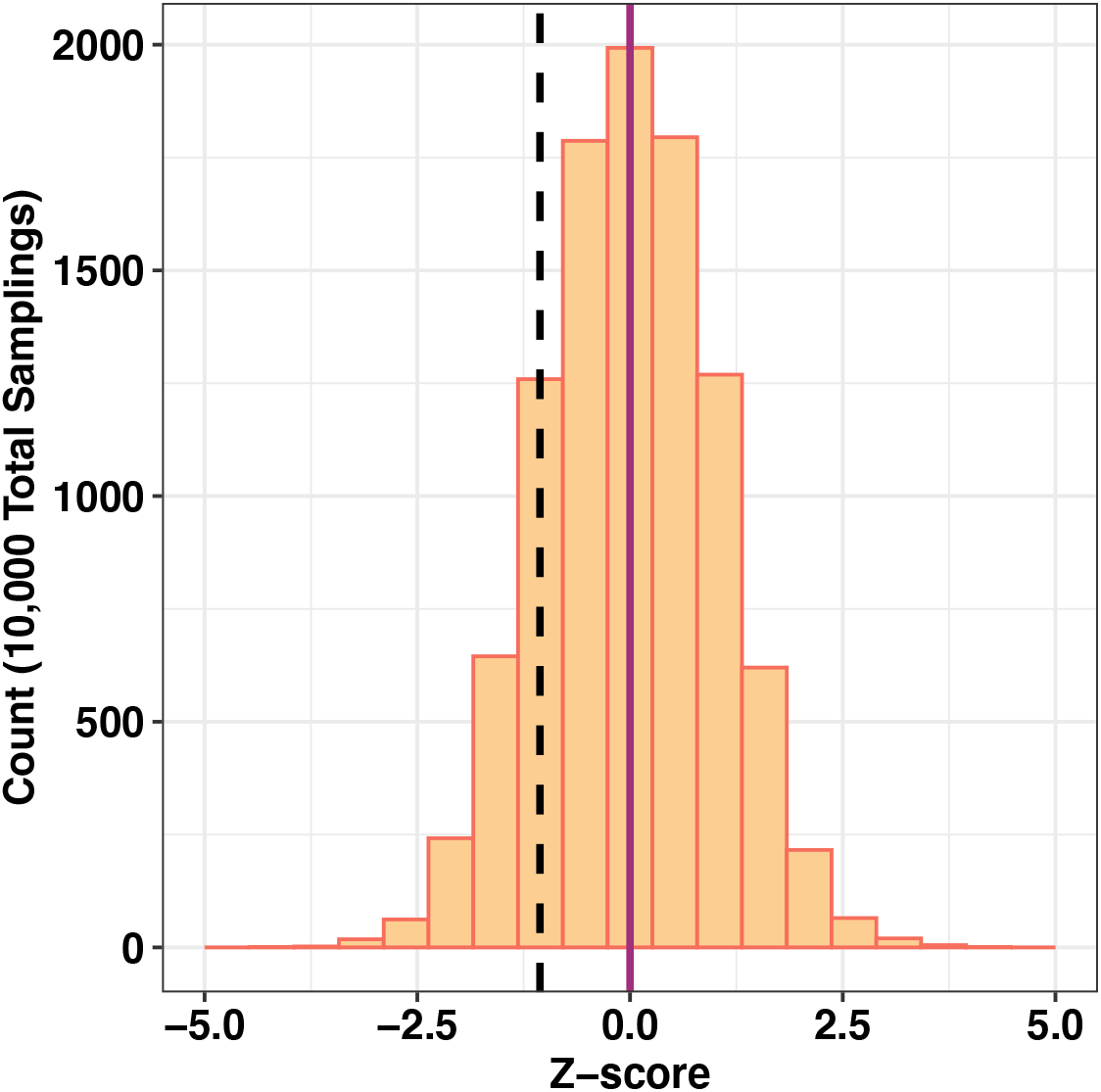
Summary of rare variant carrier association results for NAT2. The distribution of z-sores for association statistics of NAT2 rare variant burden and simvastatin use from 10,000 samples of the UKB European ancestry population. The dashed black line indicates the first observation and solid purple line labels the z-score for a null result.

### Supplementary Table Legends (provided as separate file)

**Table S1**. Prevalence of medication use phenotypes in European ancestry.

**Table S2**. Prevalence of medication use phenotypes in African ancestry.

**Table S3**. Pharmacogene coordinates.

**Table S4**. Table S4. Star (*) allele frequencies in European (EUR) and African (AFR) ancestry cohorts from the UK Biobank and EUR from the eMERGE PGx Sequencing Pilot.

**Table S5**. Metabolizer phenotype (MP) and activity score (AS) distributions in European (EUR) and African (AFR) ancestry cohorts from the UK Biobank.

**Table S6**. Association between pharmacogene activity score (AS) and metabolizer phenotype (MP) in European ancestry UKB participants.

**Table S7**. Association between pharmacogene activity score (AS) and metabolizer phenotype (MP) in African ancestry UKB participants.

**Table S8**. Association between dose of * allele and medication use for all nominally significant results in UKB EUR (shown in Table S6).

**Table S9**. Association between dose of * allele and medication use for all nominally significant results in UKB AFR (shown in Table S7).

**Table S10**. Diplotype effects for all nominally significant activity score and metabolizer phenotype associations in UKB AFR (Table S7).

## Notes

### Author Declarations

This study was exempt from Yale IRB review due to use of deidentified data. UKB data were accessed via reference number 58146 and are available to other bona fide researchers though the UKB Data Access Management System. UK Biobank has obtained Research Tissue Bank (RTB) approval from its ethics committee. Pilot sequencing data from eMERGE PGx were accessed through dbGAP (dbGaP Study Accession: phs000906.v1.p1) and are available to bone fide researchers through dbGAP Authorized Access application. Cohort-level consent was collected for participants following ethics procedures at each contributing site.

